# Value of SiPM PET in myocardial perfusion imaging using Rubidium-82

**DOI:** 10.1101/2020.04.23.20076844

**Authors:** S.S. Koenders, J.A. van Dalen, P.L. Jager, S. Knollema, JR Timmer, M. Mouden, C.H. Slump, J.D. van Dijk

**Author notes:** Corresponding author: S.S. Koenders, MSc, Isala Hospital, department of Nuclear Medicine, PO Box 10400, 8000 GK Zwolle, The Netherlands.

## Abstract

**Background:** PET scanners using silicon photomultipliers with digital readout (SiPM PET) have an improved temporal and spatial resolution compared to PET scanners using conventional photomultiplier tubes (PMT PET). However, the effect on image quality and visibility of perfusion defects in myocardial perfusion imaging (MPI) is unknown. Our aim was to determine the value of a SiPM PET scanner in MPI.

**Methods:** We prospectively included 30 patients who underwent rest and regadenoson-induced stress Rubidium-82 (Rb-82) MPI on the D690 PMT PET (GE Healthcare) and within three weeks on the Vereos SiPM PET (Philips Healthcare). Two expert readers scored the image quality and assessed the existence of possible defects. In addition, interpreter’s confidence, myocardial blood flow (MBF) and myocardial flow reserve (MFR) values were compared.

**Results:** Image quality improved (p=0.03) using the Vereos as compared to the D690. Image quality of the Vereos and the D690 was graded fair in 20% and 10%, good in 60% and 50%, and excellent in 20% and 40%, respectively. Defect interpretation and interpreter’s confidence did not differ between the D690 and the Vereos (p>0.50). There were no significant differences in rest MBF (p≥0.29), stress MBF (p≥0.11) and MFR (p≥0.51).

**Conclusion:** SiPM PET provides an improved image quality in comparison to PMT PET. Defect interpretation, interpreter’s confidence and absolute blood flow measurements were comparable between both systems. SiPM PET is therefore a reliable technique for MPI using Rb-82.

## Introduction

Myocardial perfusion imaging (MPI) using positron emission tomography (PET) is increasing in popularity over single photon emission computed tomography (SPECT) in the last years due to the increased availability of strontium-82/rubidium-82 (Rb-82) generators, higher spatial resolution and higher sensitivity and specificity [1]. In addition, PET enables quantification of myocardial blood flow (MBF), which provides valuable additional prognostic information about the extent and functional importance of possible stenosis over visual assessment [2-4].

Recently, new PET systems using silicon photomultipliers with digital readout (SiPM PET) have become available for clinical use [5-8]. In terms of system performance, the SiPM PET design results in an improved spatial and timing resolution and a relatively high count-rate capability as compared to PET scanners using conventional photomultiplier tubes (PMT PET) [5-7, 9, 10]. First oncology-PET studies showed that SiPM PET provides an improved image quality over PMT PET [10-13]. However, studies demonstrating the value of SiPM PET for MPI are still lacking. Hence, our aim was to determine the value of SiPM PET in comparison to PMT PET in MPI using Rb-82.

## Materials and methods

### Study design

We performed a prospective single center study and included 30 consecutive patients referred for MPI using PMT PET (Discovery 690, GE Healthcare; D690) with Rb-82 for the evaluation of coronary artery disease. Within three weeks after the first PET scan patients underwent a second MPI PET scan on a SiPM PET scanner (Vereos, Philips Healthcare). The local institutional ethics committee approved the study protocol and informed consent was obtained from all individual participants included in the study.

### Patient preparation and data acquisition

Patients were asked to refrain from caffeine containing beverages for at least 24 h before both scans. All patients underwent a rest scan followed by a regadenoson-induced stress scan on both scanners. First, a low-dose computed tomography (CT)-scan was performed for attenuation correction purposes. The CT scan on the D690 was performed using 0.8 s rotation time, pitch of 0.97, collimation of 32×0.625 mm, tube voltage of 120 kV and tube current of 10 mA. On the Vereos, the CT-scan was acquired using 1.5 s rotation time, pitch of 0.83, collimation of 64×0.625 mm, tube voltage of 120 kV and tube current of 22 mA. The PET acquisition protocol was similar for the D690 and Vereos. A fixed activity of 740 MBq Rb-82 was intravenously administered with a flow rate of 50 mL/min using a strontium-82/Rb-82 generator (CardioGen-82, Bracco Diagnostics Inc.) immediately followed by a seven-minute PET acquisition. Ten minutes after the first activity bolus, stress was pharmacologically induced by administering 400 µg (5 mL) regadenoson over 10 seconds. After a 5 mL saline flush (NaCl 0.9%) the second activity bolus of 740 MBq was administered followed by a seven-minute stress PET acquisition. To obtain patient’s effective radiation dose for both PET examinations we used the conversion factors of 0.00126 mSv/MBq for rest and 0.00128 mSv/MBq for stress [14, 15] resulting in a total dose of 1.9 mSv. To calculate the effective dose for the attenuation CT we used a conversion factor of 0.014 mSv/(mGy.cm) [16] resulting in 0.2 mSv for the D690 based on an average dose length product (DLP) of 11.8 mGy.cm and 0.8 mSv for the Vereos based on an average DLP of 60.5 mGy.cm.

### Image reconstruction

CT data associated to the D690 were reconstructed using an iterative reconstruction method (70% adaptive statistical iterative reconstruction algorithm, ASIR) and a slice thickness of 5 mm. CT data associated to the Vereos were reconstructed using an iterative reconstruction method (iDose level 4) and a slice thickness of 3 mm.

We applied attenuation correction to all acquired PET data after semi-automatic registration of the CT and PET using the PET data acquired between 2:30 and 7:00 minutes [17]. We reconstructed the images of the D690 with a 3D ordered subset expectation maximization (OSEM) technique using 2 iterations and 24 subsets and a Gaussian post-smoothing filter of 12 mm, as recommended by the manufacturer. The voxel size of the D690 was 3.3×3.3×3.3 mm^3^. Images of the Vereos were reconstructed with 3D OSEM using 3 iterations and 15 subsets and a Gaussian post-smoothing filter of 6 mm. The voxel size of the Vereos was 4.0×4.0×4.0 mm^3^. These Vereos settings were determined prior to our study (see appendix), based on measurements using an anthropomorphic torso phantom with a cardiac insert (model ECT/TOR/P, Data Spectrum Corp.). Intensity profiles through the cardiac insert were collected for several reconstruction settings to compare the full width at half maximum value to that of the D690. This way we obtained reconstruction settings resulting in an equivalent image resolution. For both the D690 and the Vereos data corrections were performed for decay, scatter and random coincidences, and dead time effects. We used data acquired from 2:30 to 7:00 minutes for both rest and stress scans to obtain static images. Dynamic data sets were reconstructed using 26 time frames (12×5s, 6×10s, 4×20s and 4×40s). All reconstructed images were post-processed using Corridor4DM software (v2016).

### Visual assessment

Each set of static rest and stress PET images, showing the relative perfusion, was analyzed by two expert readers in consensus. They scored the image quality, visibility of perfusion defects and interpreter’s confidence. Image quality of the static images was assessed using a four-point grading scale: 1) poor, 2) fair, 3) good and 4) excellent. Readers assessed the image quality based on myocardial count density and uniformity in well-perfused areas, signal to background noise, and shape of the left ventricle (LV). Static images were visually characterized as normal or abnormal. Abnormal scans were characterized as ischemic and/or irreversible. The interpreter’s confidence was scored as either definite or equivocal. Readers were unaware of the patient’s history or other clinical findings. Images were presented in random order and readers were blinded for the PET system.

### MBF quantification

Activity concentrations were measured in the 26 reconstructed time frames to calculate the time activity curves (TACs) for the LV, for the three vascular territories: left anterior descending (LAD), left circumflex (LCX) and right coronary (RCA) artery, and for the whole myocardium (global). The one-tissue compartment model of Lortie et al. based on a ROI methodology was used to calculate the MBF from the TACs [18]. Rest MBF was calculated without rate-pressure product correction. Furthermore, myocardial flow reserve (MFR) was calculated as the ratio between the stress and rest MBF. We categorized the global MFR values into three categories: high risk on cardiac failure with MFR <1.5, intermediate risk with MFR between1.5-2.0 and low risk with MFR >2.0 [2, 19].

Rest or stress MBF and MFR values were excluded from the comparison evaluation in case of unreliable TACs. Unreliable TACs were defined as TACs without a clear LV peak during the first-pass phase or a lack of steady state for the three vascular territories during the tissue phase, as previously described [20]. Test-retest precision was calculated as the standard deviation (SD) of the relative MBF and MFR differences, as previously defined by Kitkungvan et al. [21]. A test-retest precision ≤21% was considered acceptable [21].

### Statistical analysis

Patient-specific parameters and characteristics were determined as mean ± SD, or as percentages using SPSS (IBM SPSS Statistics for Windows, Version 24.0). Image quality, MBF and MFR measurements were compared using the Wilcoxon signed rank test. In addition, the visibility of perfusion defects and interpreter’s confidence were compared using the McNemar test. The level of statistical significance was set to 0.05 for all statistical analyses.

## Results

### Baseline characteristics

The baseline characteristics of the included patients are summarized in Table 1.

**Table 1:** Baseline characteristics of all included patients (n=30) who underwent clinically indicated Rb-82 PET MPI

| Characteristic | All patients (n=30) |
| --- | --- |
| Age (years) | 64 ± 9 |
| Male (%) | 80 |
| Weight (kg) | 87 ± 15 |
| Height (cm) | 176 ± 9 |
| BMI (kg/m<sup>2</sup>) | 28.0 ± 4.4 |
| Current smoker (%) | 13 |
| Hypertension (%) | 50 |
| Diabetes (%) | 20 |
| Dyslipidemia (%) | 40 |
| Family history (%) | 53 |

Data are presented as mean ± SD or as percentage

### Visual assessment

Image quality of the static images improved (p=0.03) using the Vereos as compared to the D690. Image quality of the D690 and the Vereos was graded fair in 20% (6/30) and 10% (3/30), good in 60% (18/30) and 50% (15/30), and excellent in 20% (6/30) and 40% (12/30), respectively, as illustrated in Fig. 1. None of the images using either the D690 or the Vereos were scored as poor. An example of the image quality for patients with high and low BMI is shown in Fig. 2.

**Figure 1.**
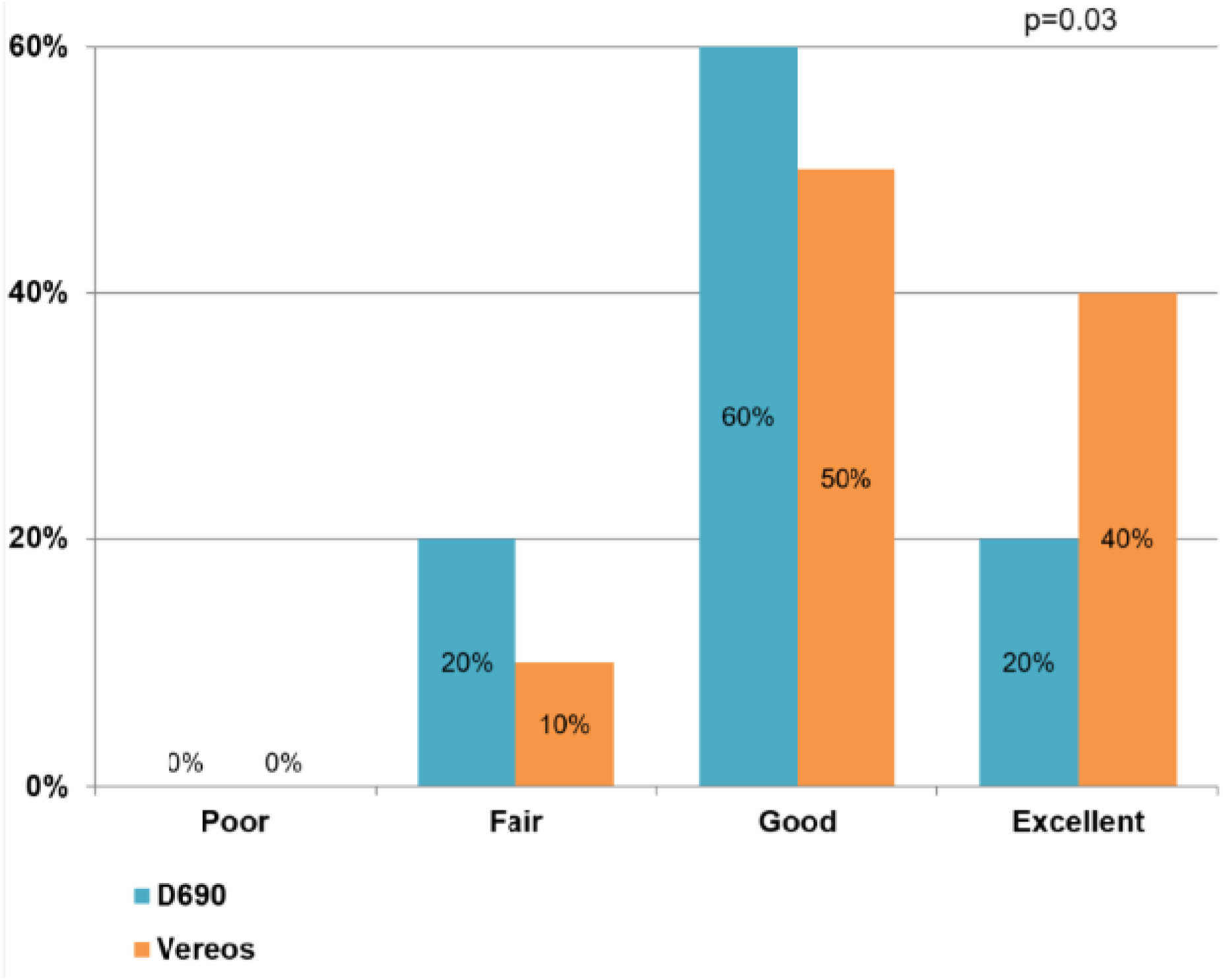

**Figure 2.**
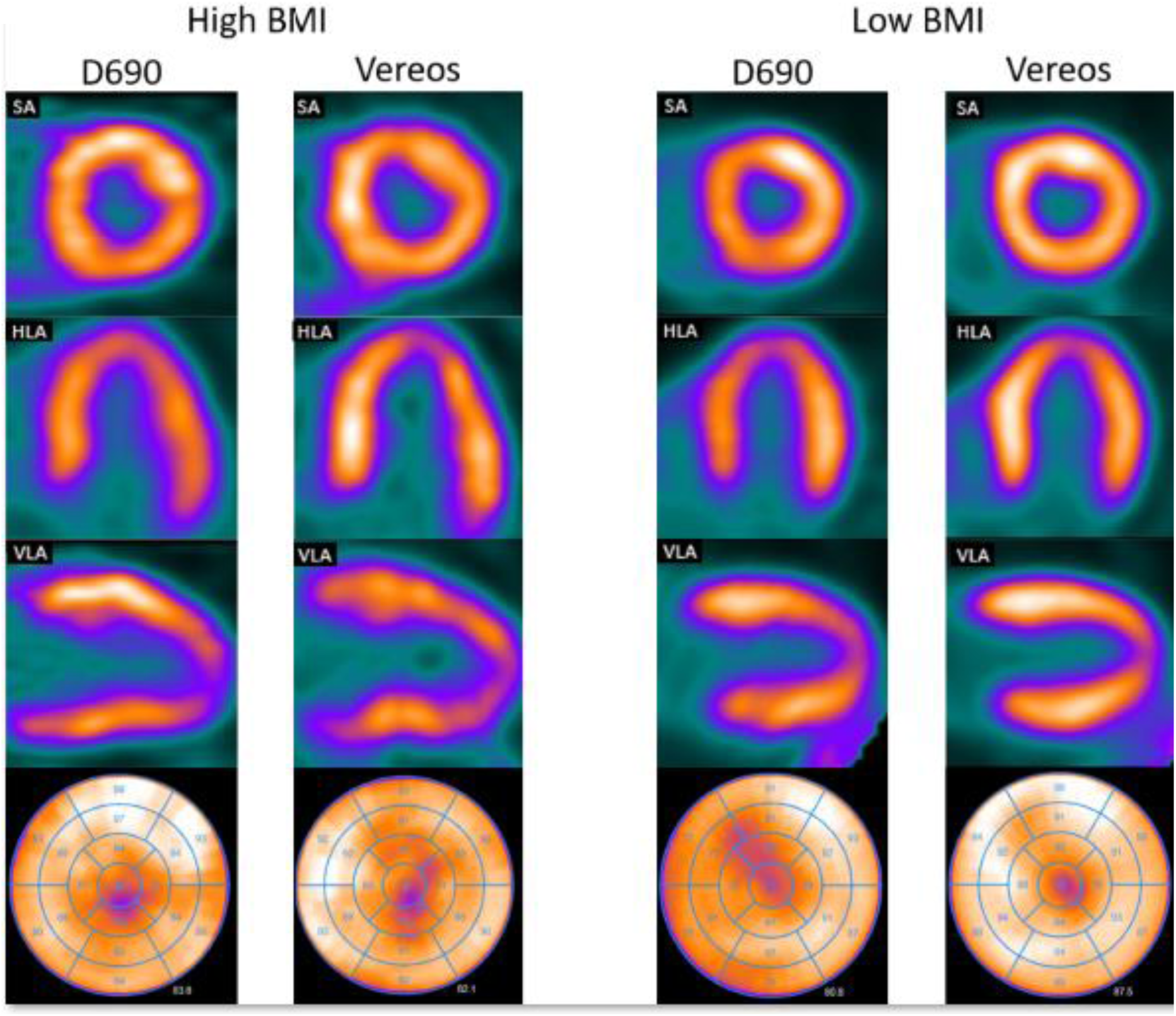

Defect interpretation did not differ in 93% (28/30) of the patient scans between the D690 and the Vereos (p=0.50). In the 7% (2/30) of patient scans where defect interpretation differed, the scans were scored as normal on the D690, whereas they were interpreted to show ischemia on the Vereos (Fig. 3 & Fig. 4). Patient scans were scored as normal in 80% (24/30) and 73% (22/30) for the D690 and the Vereos, respectively. Furthermore, 10% (3/30) and 17% (5/30) was interpreted as showing ischemia and for both PET scanners 13% (4/30) was scored as showing an irreversible defect. There was no difference in interpreter’s confidence as all scans were scored as definite.

**Figure 3.**
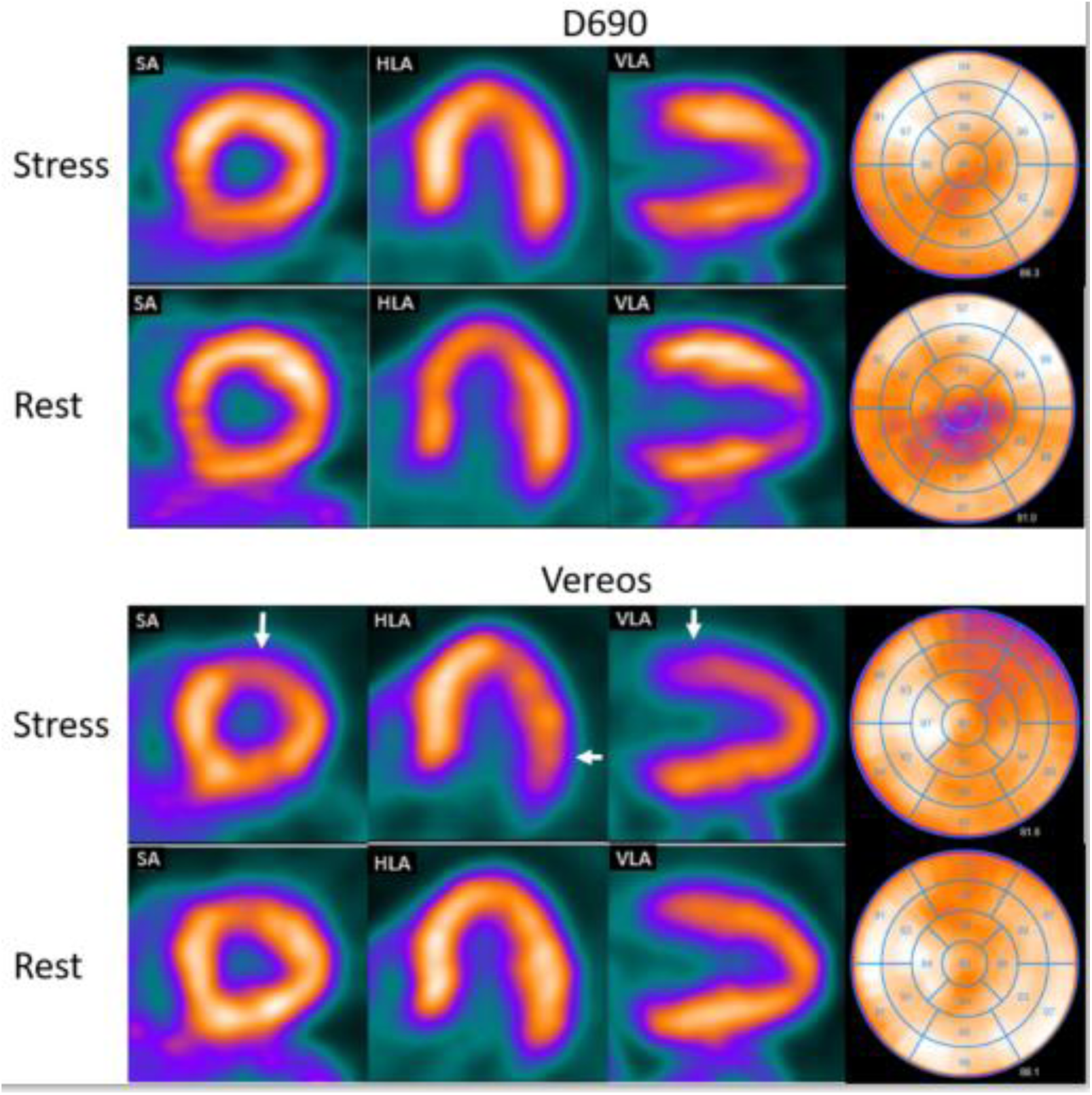

**Figure 4.**
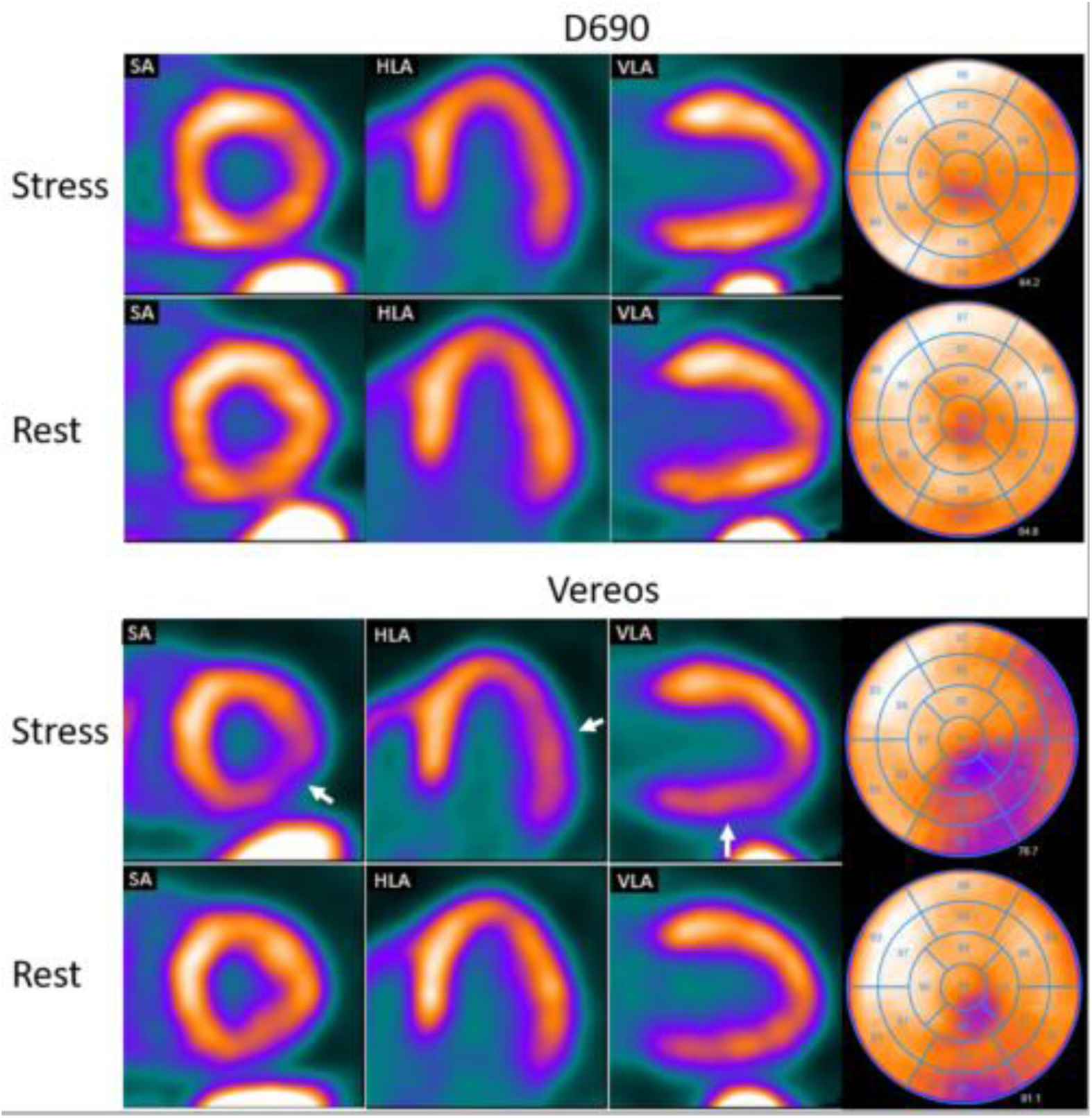

### MBF quantification

Of the 30 included patients, both rest and stress MBF values of one patient were excluded due to unreliable rest and stress TACs. Furthermore, rest MBF values of another patient and stress MBF values of four other patients were excluded due to unreliable TACs. The main reason for an unreliable TAC was no clear or absent LV peak, which would be most likely caused by a pinched vein [22]. The remaining PET scans provided a paired comparison of 28 rest MBF values, 25 stress MBF values, and 24 MFR values.

There were no significant differences in any of the vascular territories nor in the whole myocardium regarding the rest MBF (p ≥ 0.29), stress MBF (p ≥ 0.11) or MFR (p ≥ 0.51), as shown in Table 2 and Fig. 5. When categorizing the global MFR values into high, intermediate and low risk on cardiac failure, 25% (6/24) of the patients were reclassified when using the Vereos. More specifically, one patient was reclassified from intermediate risk to high risk, three from intermediate risk to low risk and two patients from low risk to intermediate risk. None of the patients were reclassified from low risk to high risk or vice versa. Moreover, test-retest precision of global rest MBF, stress MBF and MFR were 18%, 16% and 21%, respectively, and were considered to be within the previously reported test-retest precision of 21%, as shown in Fig. 6.

**Table 2:** Rest and stress MBF (mL/min/g) and MFR values calculated for the D690 and the Vereos PET scans, for the three vascular territories (LAD, LCX, and RCA) and the whole myocardium (global). No significant differences were observed between the D690 and the Vereos PET (p ≥ 0.11)

| Territory | PET scanner | Rest MBF (n=28) | Stress MBF (n=25) | MFR (n=24) |
| --- | --- | --- | --- | --- |
| LAD | D690 | 0.9 [0.7-1.1] | 2.0 [1.8-2.5] | 2.3 [2.0-2.8] |
|  | Vereos | 0.9 [0.7-1.0] | 2.2 [1.7-2.4] | 2.3 [2.0-2.7] |
| LCX | D690 | 0.9 [0.7-1.1] | 2.2 [1.9-2.5] | 2.3 [2.0-2.9] |
|  | Vereos | 0.9 [0.7-1.1] | 2.1 [1.6-2.3] | 2.3 [1.8-2.7] |
| RCA | D690 | 0.9 [0.7-1.1] | 2.2 [2.0-2.5] | 2.3 [2.0-3.0] |
|  | Vereos | 0.9 [0.7-1.2] | 2.3 [2.0-2.9] | 2.4 [1.9-3.3] |
| Global | D690 | 0.9 [0.7-1.0] | 2.0 [1.8-2.5] | 2.3 [2.0-2.9] |
|  | Vereos | 0.9 [0.7-1.0] | 2.1 [1.7-2.5] | 2.3 [2.0-2.8] |

Data are presented as median (interquartile range). LAD, left anterior descending; LCX, left circumflex; MBF, myocardial blood flow; MFR, myocardial flow reserve; and RCA, right coronary artery

**Figure 5.**
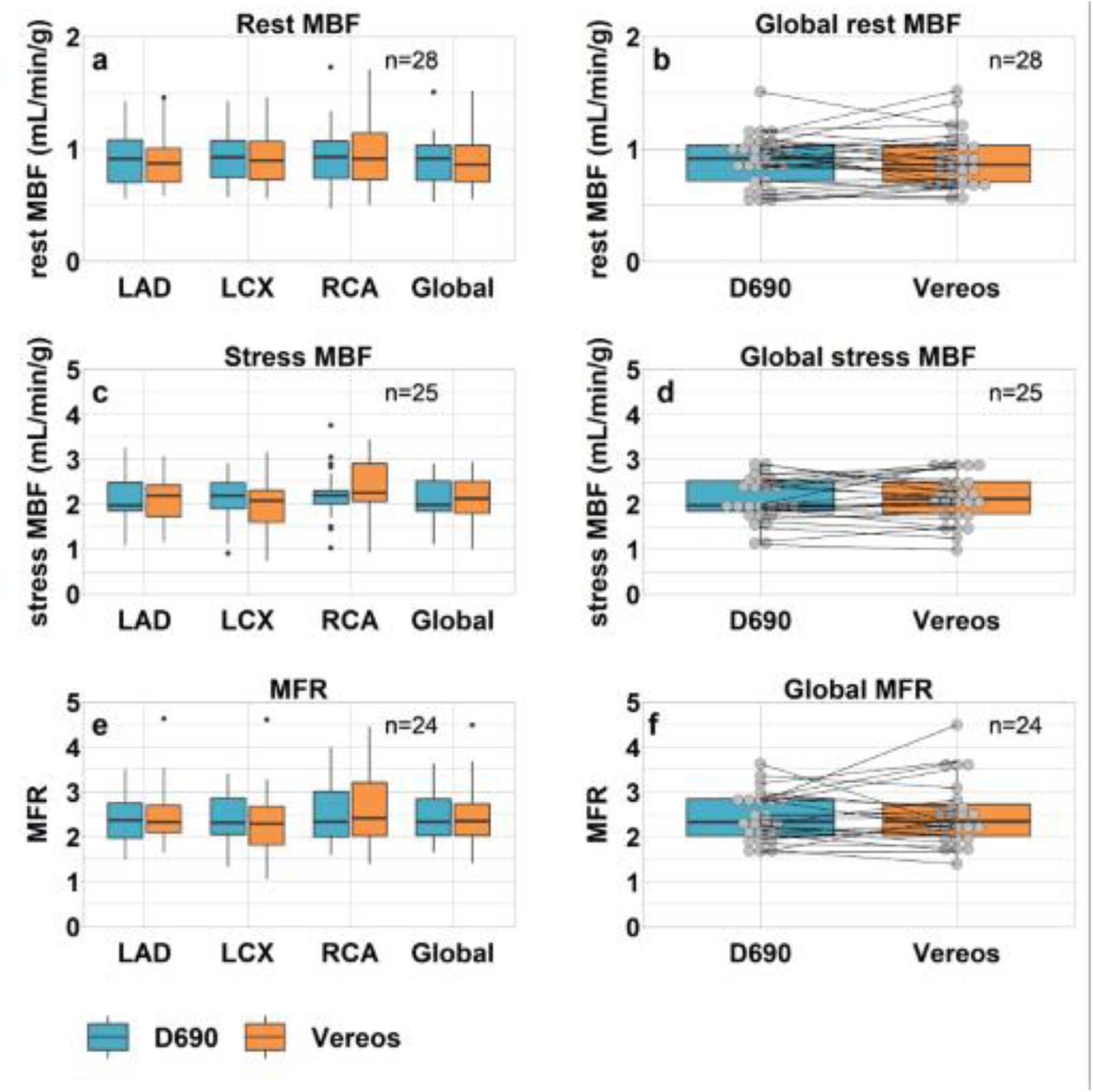

**Figure 6.**
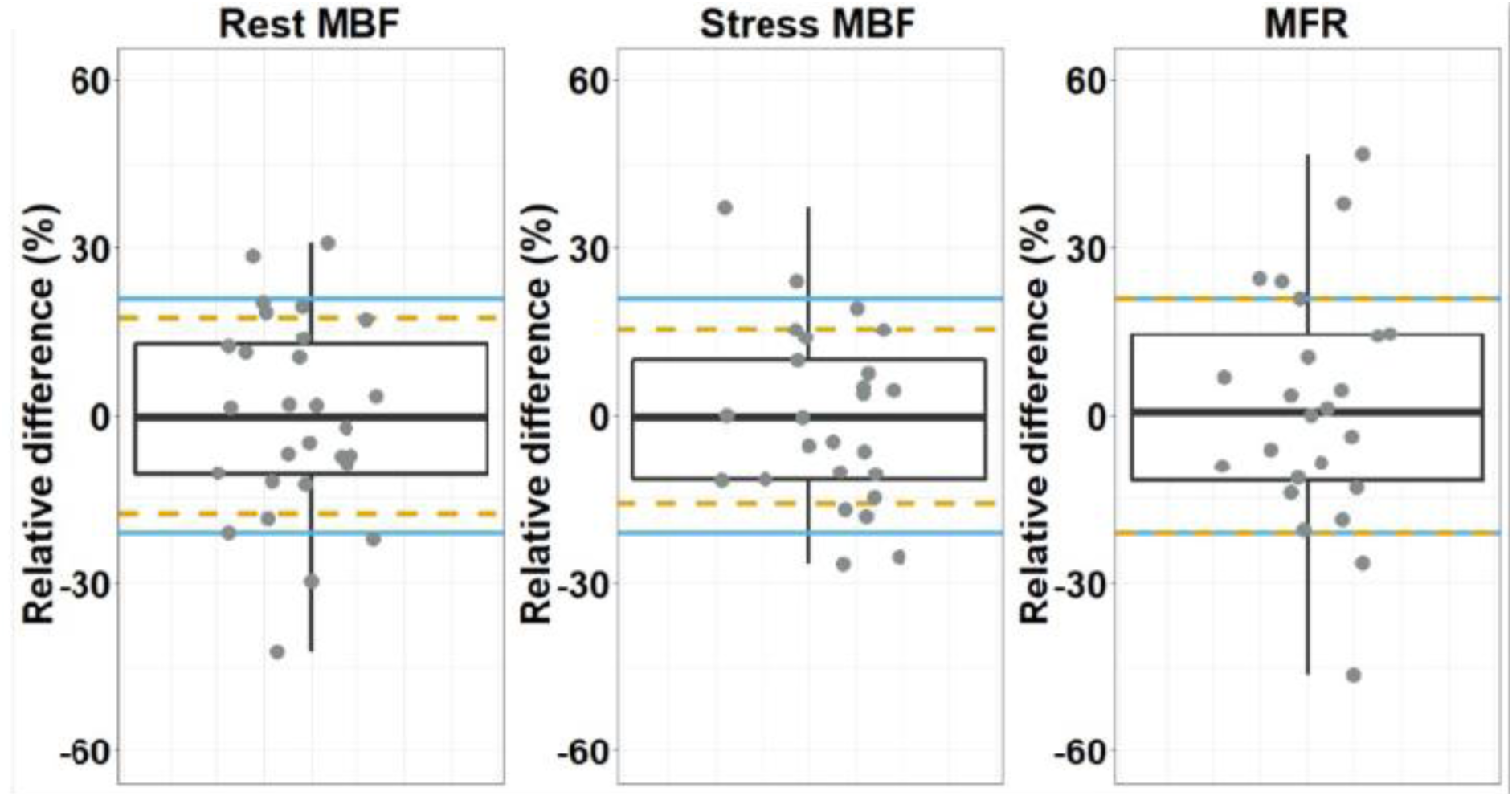

## Discussion

In this study we showed that the Vereos SiPM PET scanner provided an improved image quality for MPI using Rb-82 as compared to the D690 PET scanner using conventional photomultiplier tubes. There were no significant differences in defect interpretation or in quantitative MBF and MFR measurements.

According to previously performed phantom and patients studies [6, 10-13] SiPM PET showed an improved image quality and lesion detection for oncology patients as compared to PMT PET. It seems that these results can be generalized to cardiac imaging as shown in our study. In addition, scan interpretation might change as well when shifting from PMT PET to SiPM PET. In our population, images from two out of 30 patients showed ischemia on the Vereos PET scan, whereas these images were interpreted as normal on the D690 PET scan. Of these patients, one had no follow-up imaging and no events within the first year after the Vereos PET scan (Fig. 3). Global MFR values of this patient were 2.15 using the D690 (low risk) and 1.87 using the Vereos (intermediate risk). In the other patient (Fig. 4), a subtotal stenosis was seen in the circumflex area during coronary angiography one month after the second PET scan, corresponding to the ischemic area in the Vereos PET images. Global MFR values of this patient were 1.72 using the D690 (intermediate risk) and 1.40 using the Vereos (high risk). It is well known that perfusion defects can be introduced due to misregistration of attenuation CT and PET data [23-25]. For each scan we verified the co-registration between CT and PET data. In none of the scans a misregistration was observed.

For MBF quantification, the Vereos showed reliable MBF and MFR measurements using Rb-82. This is in line with the results of the study by Van Dijk et al. who performed a cardiac-phantom study and concluded that the D690 and the Vereos scanner showed a comparable count-rate performance for Rb-82 activities up to approximately 1000 MBq [26]. However, in our study 25% (6/24) of the patients were reclassified according to the global MFR values from intermediate risk to low/high risk or vice versa when shifting to the Vereos. The relative differences of these six patients were 38%, 15%, 10%, -14%, -19% and -21%. Although 25% seems to be a large percentage, it is possible that a patient classified as having an intermediate risk may be classified as having a low/high risk when repeating the scan and reprocessing the data, solely due to the relatively large test-retest precision in MBF and MFR measurements of typically 21% [21]. Therefore reclassification in 25% of the patients is not solely due to the differences in scanner performance. Furthermore, test-retest precision was calculated as the SD of the relative MBF and MFR differences for all patients. Therefore, by definition, approximately 68% of all test-retest values should be within the test-retest precision of 21%.

This study had several limitations that should be recognized. First, our study population was relatively small (n=30). Still, the interpreter’s confidence was scored as definite in 100% of the scans so it is unlikely that including more patients will give a significant change in the interpreter’s confidence between the Vereos and D690. However, our study did show a change in defect interpretation in 2 patients. Inclusion of more patients is necessary to find out if there is a significant change in defect interpretation and validation studies are required to determine a possible superior diagnostic performance of the Vereos SiPM PET over the D690 PMT PET. Furthermore, we found relative MBF and MFR differences within the 21% test-retest precision. It is not likely that this will change when more patients are included.

Secondly, some elements of the acquisition have to be addressed. As we used Rb-82, the positron range (5.9 mm) is rather large compared to for example ^13^N-ammonia (1.5 mm), which results in a worse image resolution compared to using other PET MPI tracers [27]. The higher spatial resolution of the SiPM PET as compared to PMT PET may therefore result in an even better image quality when using other PET tracers than Rb-82. Moreover, the injected activity used in this study is lower than generally recommended (740 MBq vs. 1110 MBq) [2], but sufficient for MBF quantification [26, 28]. In addition, we used a PMT PET scanner with a relatively high count-rate capability. Therefore, our results may not be generalizable to older PMT PET scanners as they might not be able to process the high count-rates adequately due to dead time effects [26, 28]. Inaccurate count-rate measurements can result in unreliable MBF and MFR measurements [29].

Lastly, expert readers had no access to clinical information, i.e. gender, age or calcium score when interpreting the images. Although there was no significant difference in defect interpretation, it still differed in two patients. Access to clinical information might have altered their decision-making and could have overcome this different interpretation.

### Clinical implications

The Vereos scanner was the first SiPM PET scanner available for clinical use [6] after which two other SiPM PET scanners became available, namely the Biograph Vision PET/CT (Siemens Healthineers) and the Discovery MI (GE healthcare) [7, 8]. As the performance characteristics of SiPM PET are in general better that those of PMT PET, image quality is expected to improve for all three SiPM PET scanners. Moreover, flow measurements are expected to be similar or possibly more accurate as compared to using PMT PET, provided that PMT PET has a sufficient count-rate capability [7, 8, 26, 28, 29].

Whereas the MFR was shown to be robust when using different advanced reconstruction settings or software packages, one should be cautious in the occurrence of possible systematic changes in MBF measurements [30, 31]. Furthermore, one has to be aware of a relatively large test-retest precision in MBF and MFR measurements of typically 21%. In general, a MFR<1.5 is associated with an increased risk on cardiac failure while patients with a MFR>2.0 are associated with a reduced risk on cardiac failure [2, 19]. Hence, it is possible that a patient classified as having an intermediate risk may be classified as a low/high risk patient when repeating the scan and reprocessing the data, solely due to the test-retest variation.

### New knowledge gained

The use of the Vereos SiPM system in PET Rb-82 MPI results in an improved image quality and no significant differences for visual interpretation or interpreter’s confidence in comparison to the conventional D690 PMT PET scanner. Furthermore, no significant differences were found in MBF and MFR quantification. We did find a change in visual defect interpretation in two patients. Defect interpretation may therefore differ and it could be possible that the Vereos SiPM PET system has a superior diagnostic performance over the conventional D690 PMT PET system. Additional studies with a larger patient population are required to confirm this.

## Conclusions

PET using silicon photomultipliers with digital readout is a reliable technique for MPI using Rb-82 as it provides an improved image quality and similar interpreter’s confidence, defect interpretation and absolute blood flow measurements as compared to PET using conventional photomultiplier tubes.

## Data Availability

This work was supported by a research grant of Philips Healthcare. The content of the article was solely the responsibility of the authors.

## Ethical approval

All procedures performed in studies involving human participants were in accordance with the ethical standards of the institutional and/or national research committee (METC Isala Zwolle, NL63853.075.17) and with the 1964 Helsinki declaration and its later amendments or comparable ethical standards.

## Informed consent

Informed consent was obtained from all individual participants included in the study.

## Abbreviations

CT: Computed tomography
LAD: Left anterior descending
LCX: Left circumflex
LV: Left ventricle
MBF: Myocardial blood flow
MFR: Myocardial flow reserve
MPI: Myocardial perfusion imaging
PET: Positron emission tomography
Rb-82: Rubidium-82
TAC: Time activity curve

## Appendix

### Aim

We performed a phantom study to obtain reconstruction settings for the Vereos SiPM PET resulting in similar image resolution as compared to the Discovery 690 PMT PET.

### Method

We used an anthropomorphic torso phantom with a cardiac insert (model ECT/TOR/P, Data Spectrum Corp.) which simulates myocardial uptake in the left ventricular chamber. An activity of 2590 MBq Rb-82 was injected. The phantom was scanned on the D690 PET system (GE healthcare) and on the Vereos PET system (Philips Healthcare). PET data of the phantom were reconstructed with a 3D ordered subset expectation maximization (OSEM) technique using 48 updates (2 iterations and 24 subsets) and a Gaussian post-smoothing filter of 12 mm on the D690, as recommended by the manufacturer. PET data of the phantom of the Vereos were reconstructed with OSEM using 45 updates (3 iterations and 15 subsets) and seven different Gaussian post-smoothing filters, with kernel-widths of 0, 2, 4, 6, 8, 10 and 12 mm. Intensity profiles through the images of the cardiac insert were collected for the seven reconstruction settings (Fig. 7 a-h) to compare the full width at half maximum value to that of the D690.

**Fig. 7.**
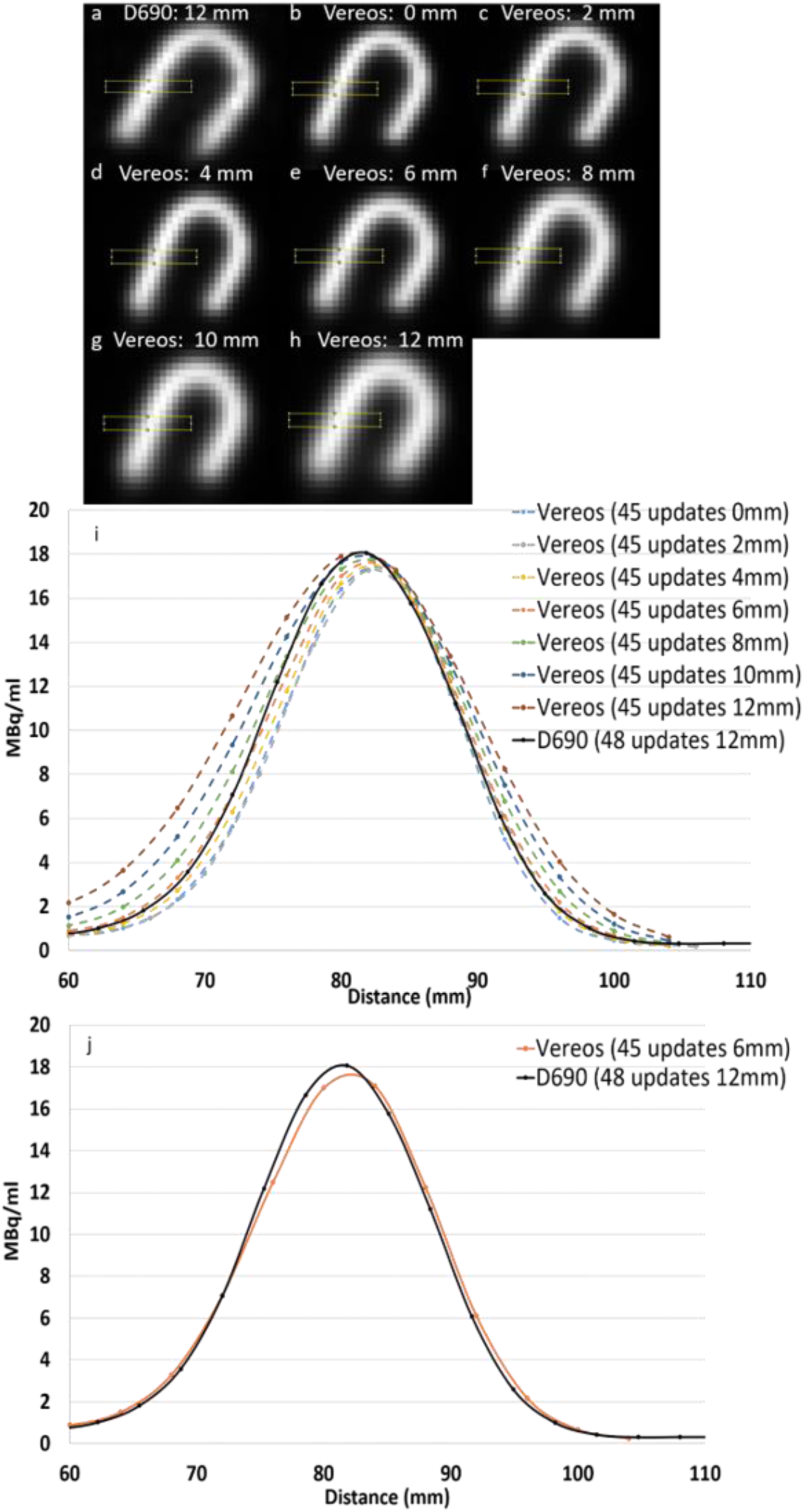
Reconstructed PET images of an Rb-82-filled Cardiac Insert phantom using the D690 (a) with 2 iterations, 24 subsets and a Gaussian post-smoothing filter of 12 mm and using the Vereos (b-h) with 3 iterations, 15 subsets and Gaussian post-smoothing filter ranging from 0-12 mm. The rectangles were used to plot intensity profiles. The width of the profile increases when increasing the filter for the Vereos reconstructions (i). The intensity profile reconstructed on the Vereos using 3 iterations, 15 subsets and a Gaussian post-smoothing filter of 6 mm is most similar to the profile using the image reconstruction of the D690 (j).

### Results

With increasing filtering, the full width at half maximum value also increases (Fig. 7i). Using a 6 mm filter in the image reconstruction of the Vereos data resulted in a similar full width at half maximum value as compared to the D690 (Fig. 7j).

### Conclusion

Reconstruction settings using 45 updates and a Gaussian post-smoothing filter of 6mm was used to reconstruct the static and dynamic images of the patients who underwent rest and regadenoson-stress PET Rubidium-82 on the Vereos.

